# Genome-wide analyses of behavioural traits biased by misreports and longitudinal changes

**DOI:** 10.1101/2020.06.15.20131284

**Authors:** Angli Xue, Longda Jiang, Zhihong Zhu, Naomi R. Wray, Peter M. Visscher, Jian Zeng, Jian Yang

## Abstract

Genome-wide association studies (GWAS) have discovered numerous genetic variants associated with human behavioural traits. However, behavioural traits are subject to misreports and longitudinal changes (MLC) which can cause biases in GWAS and follow-up analyses. Here, we demonstrate that individuals with higher disease burden in the UK Biobank (*n =* 455,607) are more likely to misreport or reduce their alcohol consumption (AC) levels, and propose a correction procedure to mitigate the MLC-induced biases. The AC GWAS signals removed by the MLC corrections are enriched in metabolic/cardiovascular traits. Almost all the previously reported negative estimates of genetic correlations between AC and common diseases become positive/non-significant after the MLC corrections. We also observe MLC biases for smoking and physical activities in the UK Biobank. Our findings provide a plausible explanation of the controversy about the effects of AC on health outcomes and a caution for future analyses of self-reported behavioural traits in biobank data.

## Introduction

Behaviours and lifestyles are modifiable risk/protective factors for common diseases in humans. In the past few decades, one of the most controversial debates in public health is on the effect of alcohol consumption (AC) on common diseases, especially cardiovascular/metabolic diseases. Large-scale meta-analyses of epidemiological studies^1,2^ on AC concluded that “no level of alcohol consumption improves health”^3^. This conclusion, however, is contradictory to the negative estimates of genetic correlation (*r*_*g*_) between AC and several diseases such as obesity^4–7^, major depressive disorder (MDD)^6,7^, Parkinson’s disease^5^ and type 2 diabetes (T2D)^5^ reported in recent genome-wide association studies (GWAS) and also contradictory to the protective effects of moderate drinking reported in observational studies^8,9^. Different hypotheses have been proposed to explain these discrepancies, including: 1) heavy AC might alter metabolism or impair nutrient absorption^10,11^, meaning that the effect is dosage-dependent; 2) people who had health problems may quit or reduce drinking, or underreport their intake level^12^; and 3) some other common explanations include confounding factors^6^ (e.g., socio-economic status and physical activities) and collider bias^13,14^. Nevertheless, to date, no study has provided in-depth investigation into the causes of the discrepancies.

In epidemiological or genetic studies, phenotypic data of behavioural and lifestyle traits are often collected from self-reported questionnaires, which are subject to misreporting (i.e., self-report biases), especially for questions related to smoking, drinking and drug use^15–18^. These behaviours are also subject to change during lifetime^19–22^, for instance in response to disease diagnosis, but data to track such longitudinal variations are rarely available. Both misreports and longitudinal changes (hereafter referred to as MLC) could change the distribution of behavioural phenotypes and thus may affect the results of both epidemiological and genetic studies.

In this study, we set out to investigate biases due to MLC in genetic analyses of self-reported behavioural traits including AC, tobacco smoking, and physical activities in the UK Biobank (UKB)^23^. The UKB includes detailed questionnaires of these behavioural traits, providing a unique resource to investigate the potential pitfalls in the analyses of self-reported phenotypes. We demonstrate that MLC could induce biases in GWAS for these traits and follow-up analyses that use summary statistics from the GWAS. We then propose a correction procedure to mitigate the MLC biases and elaborate on why some of previous studies might suffer from MLC.

## Results

### Misreports and longitudinal changes in alcohol consumption

Misreports are common in self-reported data sets^15–18^ but often overlooked in genetic analyses. Here, we focused on the analyses of AC because 1) its relationship with common diseases is controversial; 2) the data required by our investigations and corrections are available; and 3) the sample size is large (*n =* 455,607). In this study, our definition of misreports for AC includes misreporting about drinking status^24^, underreporting the AC level^15,17^, and selective recall of the question about AC level^25^, all of which might occur in the UKB. These kinds of misreports are mainly attributed to^26,27^ social desirability^28,29^ (i.e., the tendency of participants to answer questions in ways that make them viewed favourably by others) and recall bias^30,31^ (i.e., the accuracy and completeness of past events recalled by participants are influenced by subsequent events that they experienced). First, 14,488 UKB participants identified themselves as never drinkers, but combining additional data from follow-up questionnaires and medical records^32^ revealed that at least 10% of these individuals were very likely to have drinking history, e.g., previously diagnosed as having alcoholic hepatitis or alcohol use disorder (**Supplementary Note 1**). This means that classifying self-reported never drinkers as lifetime abstainers could be problematic^24^. Thus, our analyses on AC were mainly focused on current drinkers unless specified elsewhere. Second, 9,064 individuals are classified as current drinkers but reported zero consumption level, indicating possible underreporting. Third, 66,058 individuals (15.6% current drinkers) reported their alcohol intake frequency and other related questions but did not report their AC levels, suggesting a potential selective recall bias. It has been shown previously that heavy drinkers tend to be unresponsive^25^, and a high non-response rate could lead to an underestimate of the average AC level in the sample^33^. To investigate the characteristics of the suspected misreporting individuals, we examined the phenotypes of 18 common diseases in the UKB and used disease count (the number of diseases carried) as an indicator of disease burden for each participant (**Methods, Table 1**, and **Supplementary Table 1**). We observed that unresponsive individuals had a much higher mean disease count than individuals with complete responses (1.63 vs. 1.37, Welch t-test *P =* 6.35 × 10^−294^; **Table 1**). The suspected underreporting individuals (*n =* 9,064) also showed a higher mean disease count than the remaining current drinkers (1.73 vs. 1.36, Welch t-test *P =* 2.68 × 10^−8^).

**Table 1.** Alcohol consumption and health-related traits of current drinkers in different response and longitudinal change groups

| Intake level reported? | Longitudinal change group | N | Mean phenotype |  |  | Disease prevalence |  |  |  |
| --- | --- | --- | --- | --- | --- | --- | --- | --- | --- |
|  |  |  | AC | BMI | Disease count | CVD | T2D | DYSLIPID | HYPER |
| Yes | All | 358,449 | 10.67 | 27.17 | 1.372 | 0.154 | 0.051 | 0.169 | 0.184 |
|  | LESS | 152,854 | 8.56 | 27.77 | 1.506 | 0.178 | 0.072 | 0.192 | 0.211 |
|  | SAME | 134,808 | 10.82 | 26.72 | 1.274 | 0.141 | 0.038 | 0.158 | 0.167 |
|  | MORE | 68,855 | 15.11 | 26.68 | 1.266 | 0.128 | 0.031 | 0.141 | 0.154 |
| No | All | 66,058 | / | 28.28 | 1.630 | 0.177 | 0.082 | 0.181 | 0.213 |
|  | LESS | 38,799 | / | 28.70 | 1.730 | 0.194 | 0.096 | 0.195 | 0.229 |
|  | SAME | 25,055 | / | 27.70 | 1.474 | 0.153 | 0.061 | 0.159 | 0.189 |
|  | MORE | 1,599 | / | 27.40 | 1.448 | 0.138 | 0.049 | 0.147 | 0.162 |

LESS, SAME, and MORE denote the current drinkers who reduced, maintained, and increased their alcohol consumption, respectively, compared to 10 years ago. N, sample size; AC, alcohol consumption measured by units per week; BMI, body mass index; Disease count, number of common diseases affected; CVD: cardiovascular disease; T2D: type 2 diabetes; DYSLIPID: dyslipidaemia; and HYPER: hypertensive disease. The proportion of the LESS group was 42.6% in the current drinkers who reported the intake level and 58.7% in the current drinkers who did not report the intake level.

Another important source of bias is the change in drinking behaviour during the life course for reasons such as changes in health status. For instance, if people change their AC level because they are affected by a disease, such a disease ascertainment will give rise to a bias in observed or genetic relationship between AC and the disease. In the UKB, all the current drinkers (*n =* 424,507) were asked a question “compared to 10 years ago, do you drink less/the same/more nowadays?” (**Methods**), and ~62% of them reported “less” or “more”. We denote the three groups of individuals as LESS, SAME and MORE, respectively. The LESS group (*n =* 191,653) had a lower average AC level, higher disease prevalence for several common diseases, and higher mean disease count than individuals in the other two groups (**Table 1** and **Supplementary Table 1**). A follow-up question was to ask the participants to choose the reason(s) why they reduced drinking, and the available options include illness, health precaution, and financial reasons (**Table 2**). There were 15,889 individuals choosing illness/ill health or doctor’s advice as the primary reason for reducing drinking, and their mean disease count was nearly twice that of all other current drinkers (**Table 2**). In the subgroup of individuals who reported AC and had reduced drinking due to illness or doctor’s advice (*n* = 11,886), the prevalence of cardiovascular disease (CVD) was 0.411, ~2.7 times higher than that in all current drinkers (0.154), providing strong evidence of disease ascertainment of AC (**Supplementary Table 1**).

**Table 2.** Descriptive statistics of the reasons for reducing alcohol intake

| Intake level reported? | Reason of reducing | N | Mean phenotype |  |  |  |
| --- | --- | --- | --- | --- | --- | --- |
|  |  |  | AC | BMI | Disease count | EA |
| Yes | Illness or ill health | 8,555 | 7.33 | 28.53 | 2.77 | 12.81 |
|  | Doctor's advice | 3,331 | 14.72 | 29.49 | 2.58 | 12.65 |
|  | Health precaution | 48,483 | 9.98 | 27.44 | 1.54 | 14.17 |
|  | Financial reasons | 7,323 | 10.27 | 28.54 | 1.74 | 11.57 |
|  | Other reason | 68,066 | 7.54 | 27.71 | 1.28 | 13.90 |
|  | Prefer not to answer | 268 | 5.77 | 27.69 | 1.75 | 10.90 |
|  | Do not know | 16,729 | 7.28 | 27.96 | 1.35 | 13.00 |
| No | Illness or ill health | 3,541 | / | 29.28 | 2.81 | 12.35 |
|  | Doctor's advice | 462 | / | 29.99 | 2.77 | 11.90 |
|  | Health precaution | 6,767 | / | 28.26 | 1.87 | 12.86 |
|  | Financial reasons | 1,600 | / | 29.05 | 1.91 | 11.26 |
|  | Other reason | 21,362 | / | 28.62 | 1.51 | 12.88 |
|  | Prefer not to answer | 101 | / | 28.23 | 2.10 | 9.56 |
|  | Do not know | 4,934 | / | 28.79 | 1.53 | 11.89 |

N, sample size; AC, alcohol consumption (units per week); BMI, body mass index; Disease count, number of common diseases affected; EA, educational attainment (years of schooling). A total of 132 individuals did not have records for their reasons for reducing their alcohol intake.

### Biases in GWAS for alcohol consumption due to MLC

We conducted GWAS analyses for AC with and without correcting for MLC. The MLC corrections included excluding individuals who might underreport AC level, excluding individuals who reduced drinking due to illness/doctor’s advice, and adjusting the mean and variance difference in three longitudinal change groups (**Methods** and **Supplementary Figures 1-2**). There were 53 and 47 independently genome-wide significant loci (*P*_GWAS_ < 5 × 10^−8^) before and after the corrections, respectively (**Supplementary Table 2** and **Supplementary Figure 3**). We identified 16 loci that became non-significant after the corrections (*P*_GWAS_ ≥ 5 × 10^−8^, **Supplementary Table 2**). By searching the top associated SNPs at these loci in an online database PheWAS^34^ (**URLs**), we found that 44.9% of associated phenotypes (*P*_PheWAS_ < 5 × 10^−8^) were metabolic/cardiovascular traits such as body mass index (BMI), triglyceride (TC), and coronary artery disease (CAD) (**Figure 1**). We showed by a down-sampling analysis that the number of loci that became non-significant after the MLC corrections was significantly larger than that expected from a loss of sample size, and that 10 loci that became genome-wide significant after the MLC corrections were likely to be masked by MLC in the uncorrected GWAS (**Methods, Supplementary Table 2-3**). These results were in line with the simulation results (**Methods** and **Supplementary Note 2**) that MLC could reduce the power to detect true signals and induce spurious signals due to disease ascertainment (**Supplementary Figures 3-9**).

**Figure 1.**
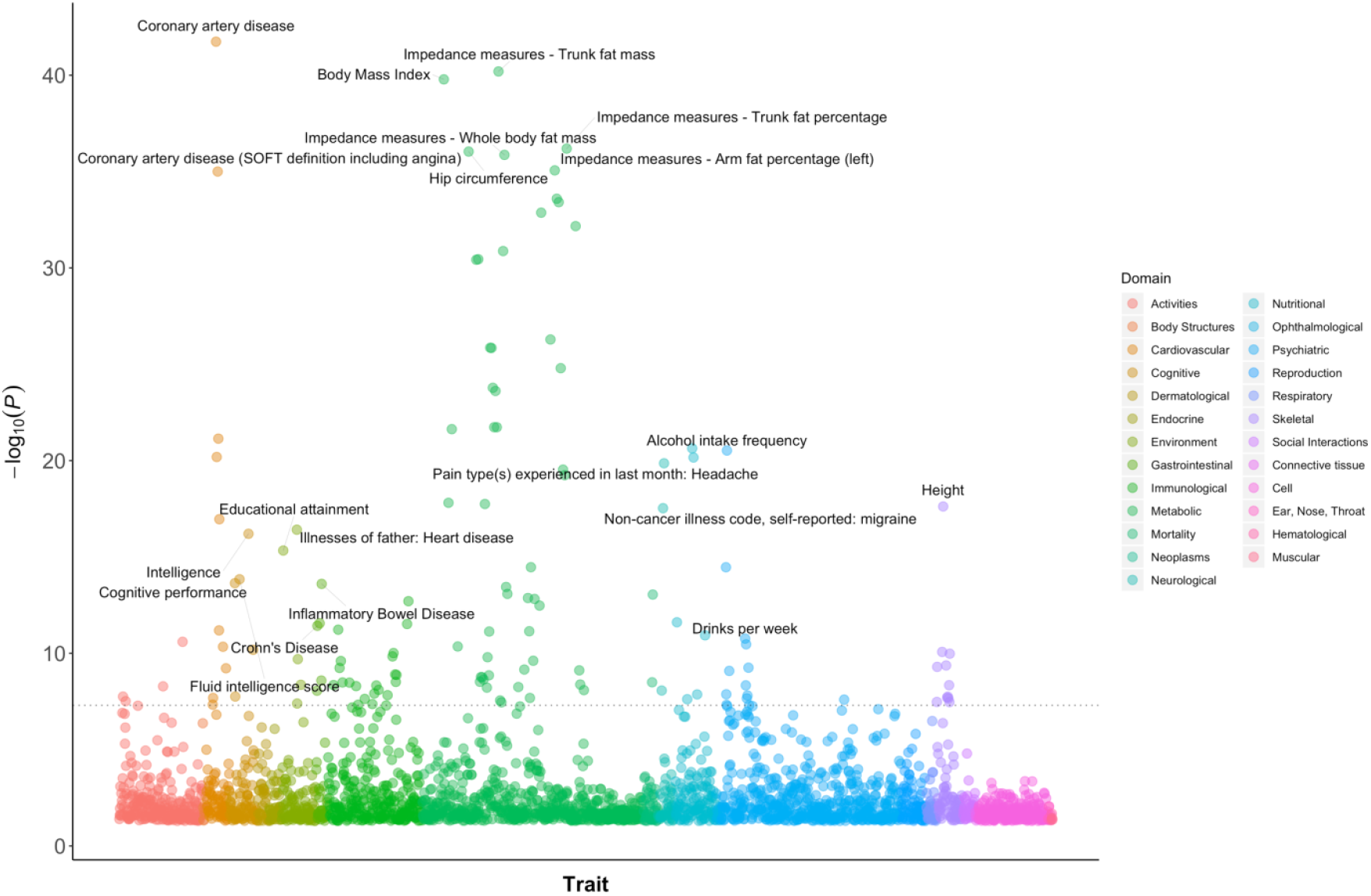
PheWAS results for the 16 AC GWAS signals that became non-significant after the MLC corrections. The colour denotes the domain of the associated trait. There were 136 traits associated with the 16 SNPs with *P* < 5 × 10^−8^, and 61 (44.9%) of them were metabolic/cardiovascular traits.

### Estimates of genetic correlation biased by MLC

Biases in GWAS data due to MLC are expected to carry over to follow-up analyses using summary statistics of the GWAS, such as the genetic correlation (*r*_*g*_) analysis. To demonstrate such biases, we estimated *r*_*g*_ between AC and 18 common diseases in the UKB by the bivariate LD score regression^35^ (LDSC) (**Methods**) using AC GWAS data from each of the three longitudinal change groups or the whole sample. Before the MLC corrections, we observed substantially differences between 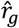 (between AC and diseases) estimated using AC GWAS data from the LESS, SAME and MORE groups (**Table 1**). We also estimated *r*_*g*_ between different AC GWAS data sets (**Supplementary Tables 5-6**, and **Supplementary Figure 10**) and found that the 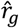 between AC in the LESS and MORE groups was significantly different from 1 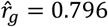, standard error (s.e.) *=* 0.074). All these results suggest that there was heterogeneity between AC data from the three longitudinal change groups. The heterogeneity was also demonstrated in an additional analysis where we estimated *r*_*g*_ between AC (using data from the UKB) and 234 traits (using data from LD-Hub^36^) and found that the 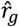 using AC GWAS data from the LESS group were substantially different from those using AC GWAS data from the MORE group, with more than half of the 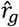 (143/234) in the opposite direction between the two groups (**Supplementary Figure 11**). Notably, after the MLC corrections, the 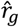 between all pairwise AC GWAS data sets were close to 1 (ranging from 0.91 to 0.99; **Supplementary Table 6**).

In the *r*_*g*_ analysis using AC GWAS data from the whole sample without the MLC corrections, AC showed nominally significant (p < 0.05) negative 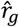 with 3 diseases (i.e., T2D, hypertensive disease and iron deficiency anemias) (**Figure 2** and **Supplementary Table 4**). Negative estimates of *r*_*g*_ between AC and diseases have also been reported in the literature. For instance, Clarke et al.^4^ show a negative 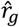 between AC and obesity, and Liu et al.^5^ show that AC is negatively genetically correlated with several common diseases including Parkinson’s disease, obesity, and T2D. However, after the MLC corrections, AC showed nominally significant 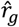 with 8 diseases, all of which were positive (**Figure 2**). These results imply that the negative estimates of *r*_*g*_ between AC and diseases from the analyses without the MLC corrections (including those in prior works) were caused by disease ascertainment. Nevertheless, this conclusion is not definitive because the ground truth is unknown in real data analysis. Hence, we sought to verify it by simulation (**Methods** and **Supplementary Note 2**), and the results showed that the estimated SNP effect correlation 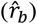 between a simulated phenotype and disease gradually changed to the opposite direction as the strength of disease ascertainment increased (**Supplementary Note 2** and **Supplementary Figures 5** and **7**), supporting our conclusion. The simulation results also showed that after MLC corrections, *r*_*b*_ was slightly underestimated but with no bias in direction in the presence of disease ascertainment (**Supplementary Figure 9**).

**Figure 2.**
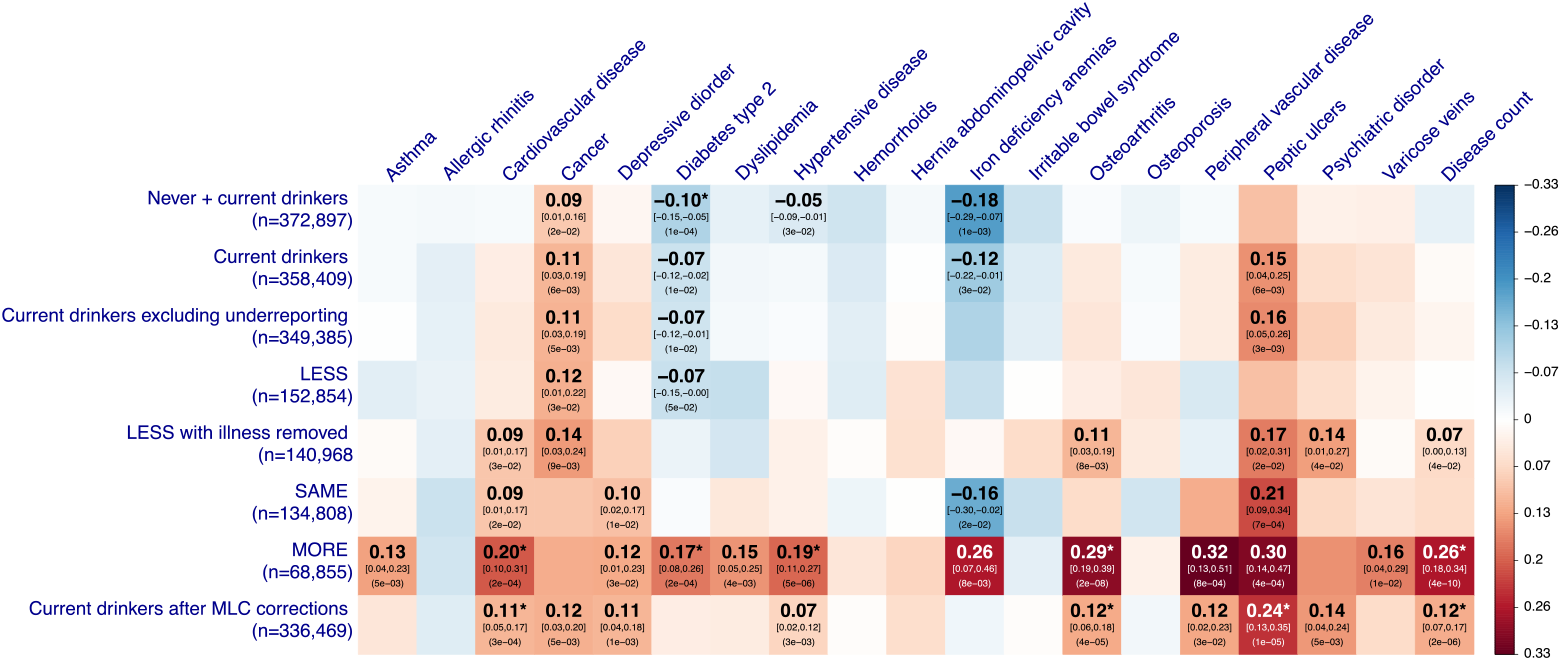
Estimates of genetic correlation between AC and common diseases in the UKB. The rows denote 8 GWAS summary data sets for AC with the sample size labelled in the bracket. The columns are 18 common diseases and disease count. The nominal significant effects (*P* < 0.05) are labelled with 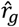 [95% confidence interval] (*P*-value), and the significant effects after multiple testing correction (*P* < 0.05/152) are labelled with an additional asterisk. “Current drinkers excluding underreporting” represents current drinkers excluding 9,064 individuals who likely underreported their AC levels. LESS, SAME, and MORE represent current drinkers whose AC levels were reduced, maintained the same, and increased, respectively, compared to 10 years ago. “LESS with illness removed” represents the LESS group excluding the participants who reduced their AC intake level due to illness or doctor’s advice.

**Figure 3.**
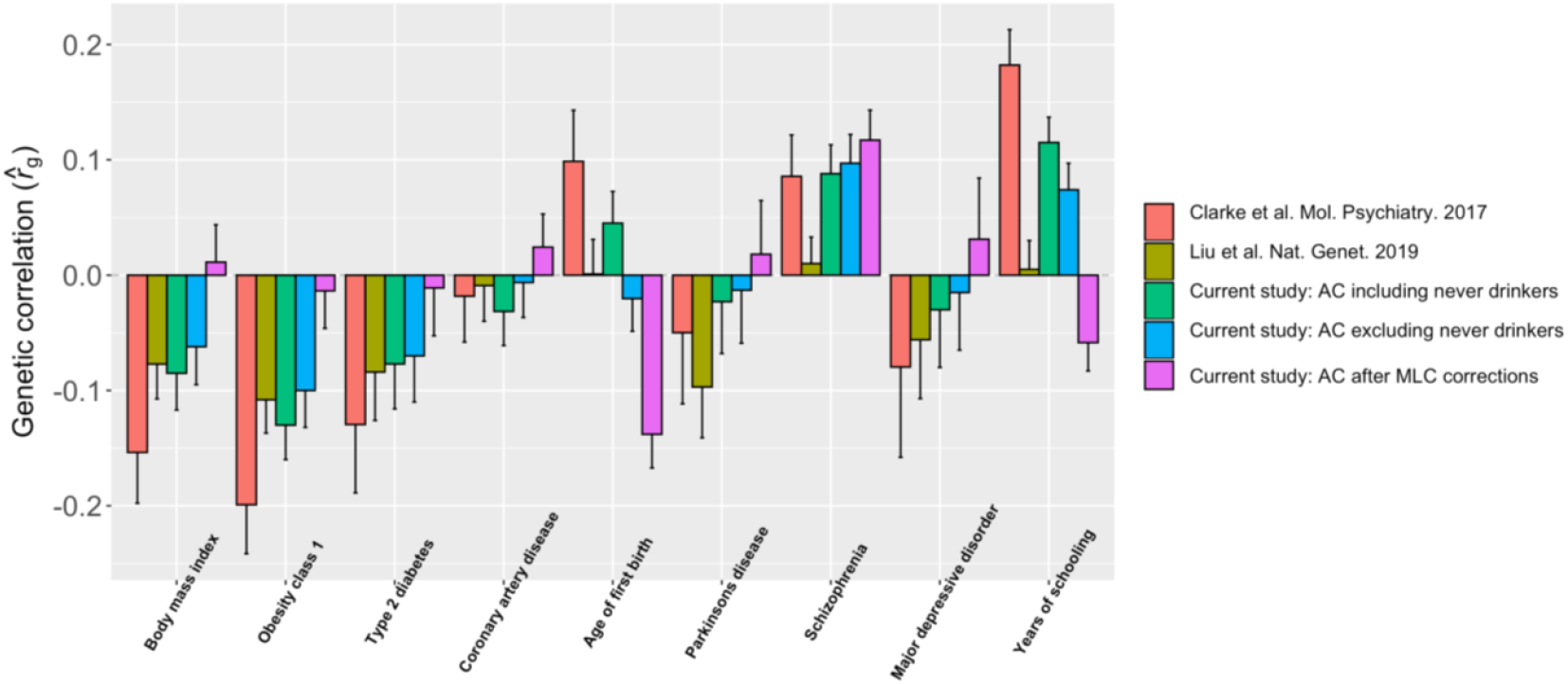
Estimates of genetic correlation between AC and complex traits using data from the UKB and other published studies. Genetic correlation was estimated by the bivariate-LDSC in LD Hub. The y-axis shows the estimate of *r*_g_, and the x-axis shows different complex traits. The error bars denote the standard errors of the estimates. The results using the summary statistics from our analysis were compared to those from Clarke et al. (2017, Molecular Psychiatry)^4^, who used self-reported AC from the interim release of the UKB data, and Liu et al. (2019, Nature Genetics)^5^, a meta-analysis that included the full release of the UKB data.

Socio-economic status (SES) has been shown to affect people’s alcohol use and health outcomes, and several studies have shown that people with higher SES tend to have higher AC levels and lower disease risks than people with lower SES^37,38^. Clarke et al.^4^ and Liu et al.^5^ show positive 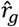 between AC and educational attainment (EA). We observed a similar estimate in our study (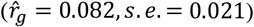 before the MLC corrections, but the estimate became non-significant after the MLC corrections (**Figure 3**), likely because MLC are associated with EA. For example, the mean years of schooling of individuals who reduced AC due to illness/doctor advice (12.76, *s. e. m. =* 0.05) was significantly lower than that of the remaining current drinkers (14.23, *s. e. m. =* 0.01). We also included household income (HI) and social deprivation (SD) in the analysis (**Methods**), and our results showed that 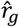 between AC and HI or SD could also be biased by MLC (**Supplementary**

### Estimates of causal effect biased by MLC

Mendelian randomization (MR) is a method that uses genetic variants as instrumental variables to infer causal relationship between exposure and outcome^39,40^. As the MR analysis relies on GWAS data, it may also be affected by the MLC biases as described above. We used BMI in the UKB as an example to demonstrate the performance of MR in the presence of MLC, based on several commonly used MR methods including IVW (inverse variance weighted)^41^, MR-Egger^42^, GSMR^43^, weighted median^44^. While the estimates from weighted median, MR-Egger and GSMR were all positive and consistent across all the analyses with or without the MLC corrections, the estimates from IVW, simple median^44^ and MR-PRESSO^45^ seemed to be sensitive to MLC with some of them being negative (**Figure 4**). The negative estimates from the analyses without the MLC corrections were likely to be driven by the 16 loci that were removed by the MLC corrections (note that the mean per-SNP MR estimate at the 16 loci was −0.077). GSMR provided the most robust estimates among the MR methods tested (**Figure 4** and **Supplementary Table 10**) because it has a step (called HEIDI-outlier filtering) that can identify and remove some of the genetic instruments whose effects were biased by pleiotropy/confounding, diminishing the bias in the estimated causal effects (**Supplementary Figure 14**). Notably, after the MLC corrections of AC, the estimates from all the MR methods were largely consistent (**Supplementary Table 10**). We also ran the GSMR analysis in a reverse direction without the MLC corrections and found significant negative effect of BMI on AC (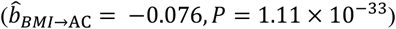 (**Supplementary Table 11**), consistent with the observation above that high BMI might be one of reasons to reduce AC (**Table 1**).

**Figure 4.**
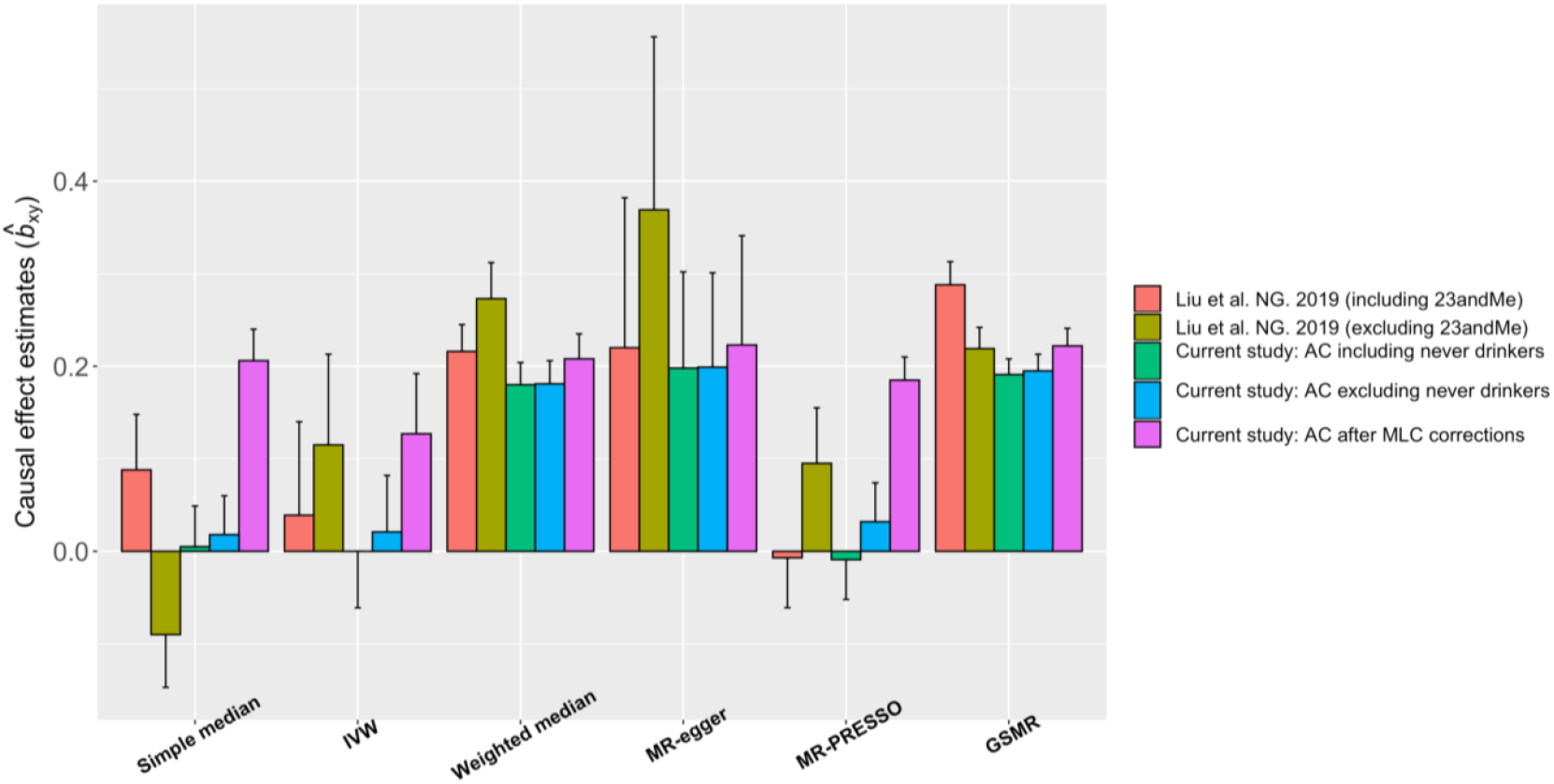
Estimates of causal effect of AC on BMI using different MR methods. The colour denotes five different GWAS summary data sets for AC. The y-axis shows the estimate of causal effects, and the x-axis shows different MR methods. The error bars denote the standard errors of the estimates.

In addition to the UKB data, we also analysed GWAS summary data for AC from Liu et al.^5^ with a sample size of ~1 million consisting of ~42.9% of the sample from 23andMe and ~33.0% from the UKB (**Methods**). The results were similar to those from the analyses above using the AC GWAS data from the UKB only (**Supplementary Table 10** and **Supplementary Figure 15**), which implies that MLC may not be UKB-specific but also exist in other data sets because otherwise the biases would be smaller in this analysis given only one-third of the AC data were from the UKB. We further confirmed the biases in MR analyses from MLC by simulation (**Supplementary Figures 6** and **8**), and demonstrated that the estimates of causal effects from MR were nearly unbiased after the MLC corrections (**Supplementary Figure 9**).

### The J-shaped relationship between AC and CVD

In epidemiological studies, there are debates about whether moderate drinking is protective against CVD because of an observed J-shaped relationship between AC and CVD^2,19,46^. We showed that the moderate drinking group (0 < AC ≤ 25 gram/week), often used as the reference group to compute the effect (odds ratio, OR) of AC on disease risk, was enriched with individuals from the LESS group which had a higher CVD incidence than the SAME and MORE groups (**Supplementary Figure 12**). This could result in a higher CVD incidence in the reference group than average, leading to a J-shaped relationship between AC and CVD (**Supplementary Figure 13A**). Although the J-shaped relationship between AC and CVD did not change much after MLC corrections (**Supplementary Figure 13B**), it became monotonically increasing after excluding the LESS group from the reference (**Supplementary Figure 13C**). Polygenic predictor of AC showed no evidence for any protective effect of moderate drinking against CVD (**Supplementary Figure 13D**), consistent with the result from a previous study^46^. Our results indicated that the J-shaped relationship between AC and CVD observed in epidemiological studies might be driven by disease ascertainment (**Supplementary Note 3**).

### Biases from MLC in other self-reported behavioural traits

Self-reported smoking data in the UKB is also likely to suffer from MLC. Similar to that for AC, all the current smokers were asked “Compared to 10 years ago do you smoke less/the same/more nowadays?”. We partitioned the current smokers (*n =* 32,801) into the LESS, SAME, and MORE groups (**Supplementary Note 4**). The LESS group had a higher disease count (1.69, *s. e. m. =* 0.01) than the SAME group (1.56, *s. e. m. =* 0.01) but a lower disease count than the MORE group (1.84, *s. e. m. =* 0.03) (**Supplementary Table 12**); these results were different from those observed in AC. In the LESS group, individuals who had reduced CPD because of illness/doctor’s advice had nearly twice the disease count (2.73, *s. e. m. =* 0.03) compared to the mean in the entire sample (1.45, *s. e. m. =* 0.002) or all current smokers (1.66, *s. e. m. =* 0.01), indicating that smoking intensity was also ascertained by disease burden. However, unlike AC, the 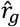 between CPD and common diseases were mostly consistent across the LESS, SAME and MORE groups (**Supplementary Figure 16**), and there were negligible differences between the 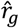 estimated using the CPD GWAS data of the whole sample before and after correcting for MLC (**Methods**; **Supplementary Table 13** and **Supplementary Figure 16**). These results are partly due to the difference in longitudinal change pattern between AC and CPD in the UKB (**Supplementary Tables 1** and **14**). For example, the mean AC in the subgroup of individuals who reduced AC due to illness (7.33 units/week) was lower than that in the entire LESS group (8.56 units/week) but with a significantly higher mean disease count, resulting in a negative correlation between AC and diseases (**Table 2** and **Supplementary Table 12**), whereas the mean CPD in the subgroup due to illness (12.28 cigarettes/day) was higher than that in the entire LESS group (11.70 cigarettes/day) (**Supplementary Table 12**).

Finally, we investigated physical activities (PA) in the UKB. The PA traits included self-reported METT (Metabolic Equivalent Task in Total) scores, IPAQ (International Physical Activity Questionnaires - short form, **URLs**), and overall acceleration average (OAA, measured by wrist-worn accelerometers). IPAQ is a derived categorical trait (low, moderate, and high) that utilizes information from the METT and its three subsets: walking, moderate, and vigorous activities (**Methods, URLs**). We first estimated *r*_*g*_ between METT, IPAQ and OAA and between METT from the three IPAQ subgroups (**Supplementary Figure 17**). We found a significant genetic heterogeneity between METT and IPAQ 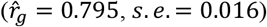 and a small genetic overlap of either METT or IPAQ with OAA (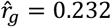 with *s. e. =* 0.037 for METT and 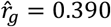 with *s. e. =* 0.034 for IPAQ). We then estimated *r*_*g*_ between PA and 18 common diseases. While the 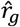 of IPAQ and OAA with the diseases were mostly negative, METT showed positive 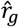 with most diseases (**Supplementary Table 15** and **Supplementary Figure 18**). We also found that the 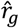 of METT from the low IPAQ subgroup with the diseases were highly consistent with those of IPAQ and OAA but mostly in the opposite direction to those of METT from the moderate and high IPAQ subgroups (**Supplementary Figure 18**), indicating potential biases in METT from the moderate and high IPAQ subgroups, in line with the finding from a previous study^47^. In addition, the phenotypic correlation between the first and third assessment (*n =* 11,484) of METT was only 0.431, implying substantial longitudinal changes. Unfortunately, these changes are undocumented for the majority of UKB participants. Given that the 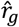 of IPAQ with diseases were highly consistent (in direction) with those of OAA (**Supplementary Figure 18**), IPAQ appears to be a more robust indicator for PA than METT.

## Discussion

In this study, we raised concerns that genetic analyses of human behavioural traits could be biased by misreports and longitudinal changes. AC in the UKB was used as the main example to demonstrate the detrimental effects of MLC on several genetic analyses commonly used to identify variant-trait associations or estimate genetic or causative relationship between traits. Our results showed that disease ascertainment was likely to be a main cause of the MLC biases, which can be largely corrected for using additional information (e.g., intake frequency and medical records) and coarse longitudinal data (e.g., self-reported longitudinal change direction). Our results also showed that the MLC corrections procedure proposed in this study added value to the routine quality controls in GWAS for behavioural traits like AC. Additionally, biases due to longitudinal changes appeared to be larger than that due to misreports, because the longitudinal changes were observed in more than half of the participants, while misreports only accounted for 10~20% of the UKB sample (at least for those we have identified thus far), as verified in our simulations (**Supplementary Note 2** and **Supplementary Figures 5-8**).

Our findings provide a plausible explanation for the long-standing controversy about the effects of AC on health outcomes in genetic^4,5^ and epidemiological studies^2,3,19^. While it seems that most inconsistent estimates in previous studies were due to MLC, there are several reasons why some studies suffered from stronger biases than others. First, the average AC level varied across datasets (from 2.9 to 19.3 units/week across 24 studies)^5^, suggesting heterogeneity in drinking behaviours among different regions/populations. Second, since we have demonstrated that biases from MLC were mainly attributable to disease ascertainment, difference in disease prevalence between populations may lead to different patterns of MLC. Third, as MLC are related to other factors, such as SES, adjusting those factors for AC may offset some of the biases in one way or another. Last but not least, MLC could vary for different age or sex groups. For instance, disease ascertainment is expected to have a larger influence in middle-aged populations than in younger populations because younger populations are less likely to be affected by common diseases investigated in this study^48^. This is supported by the observation that the older UKB participants had a higher mean disease count with a higher proportion of them reducing AC due to illness/doctor’s advice (**Supplementary Figure 19**). We also observed differences in the male/female ratio between the three groups (0.551 in the LESS group, 0.488 in the SAME group, and 0.373 in the MORE group were male). Note that we fitted age and sex as covariates in GWAS so the biases due to age or sex are likely to be limited.

Our study certainly has limitations as it is almost impossible to correct for all the biases with limited availability of relevant data. First, the 9,064 individuals who were suspected to underreport their AC are very likely to be only a subset of all the underreporting individuals. Thus, more effective methods are needed to identify the remaining underreporting individuals. Second, ~58% of the individuals with reduced alcohol intake reported that the reduction was due to “other reasons” or “do not know” in the survey (**Table 2**). If some of these individuals reduced AC also because of illness, then not taking that into account could retain a bias in the analysis. Third, some participants may have misreported their longitudinal changes, leading to an incorrect classification of longitudinal change groups. Fourth, the coarse longitudinal change information itself is cross-sectional (10 years before the time point of the first assessment), meaning that some of the changes occurred beyond the time frame might not be accounted for in this study. Last but not the least, 15% of the current drinkers who did not report their AC level were removed from the analysis. One solution, as implemented in a previous study^49^, is to impute the missing values based on intake frequency and gender. However, 99.8% of the nonresponsive individuals in the UKB are occasional drinkers while this proportion is only 9.4% for the responsive individuals, which might lead to a systematic error between the observed and imputed data sets.

In conclusion, we advise awareness of the pitfalls when analysing data on behavioural traits in biobank data sets such as the UKB. Misreports and longitudinal changes of behavioural traits by disease ascertainment could create biases and thereby induce spurious signals and a loss of power in GWAS. Biases in GWAS summary statistics due to MLC could further lead to biased estimates in follow-up analyses such as genetic correlation and Mendelian randomization. As more biobank data sets have become accessible, it is important to identify, investigate and correct for these biases in all kinds of behavioural traits including smoking, drinking, diet, physical activity, sleep, and self-rated status. A longitudinal study of 1 million individuals for several decades seems impractical at present, but we have shown that the biases in AC can be largely corrected for by phenotypic QC and longitudinal adjustment when additional phenotype information (intake frequency, medical records, longitudinal change and reasons, etc.) are available. Questionnaires on lifetime use may provide more accurate estimates of the effects of behaviours on health outcomes at a much lower cost than a longitudinal follow-up study. Researchers should be more careful regarding these biases when conducting analyses for behavioural and other modifiable traits using biobank data sets with self-reported records.

## Methods

### Phenotypic data and quality controls

We obtained behavioural and disease traits from the UK Biobank (UKB) data^50^. UK Biobank has approval from the North West Multicentre Research Ethics Committee (MREC), and informed consent has been obtained from all participants. There were 455,607 individuals of European ancestry with complete information on sex, age and principal components (PCs). The self-reported drinking status classes (data-field ID: 20117) were: never drinkers (*n* = 14,488), previous drinkers (*n* = 15,912), current drinkers (*n* = 424,507), and unknown (446 participants preferred not to answer and 254 provided no response). We removed “former drinkers” from all analyses in this study, considering the occurrence of the “sick quitter phenomenon”^51^. Among the 424,507 current drinkers, there were 358,449 individuals who reported their intake level. The AC level was summed up as a weekly total intake score (units/week) of all the alcoholic drink subtypes including beer plus cider, red wine, champagne plus white wine, spirits, and fortified wine. The mean of AC was 10.67 units per week (*s*.*d*. = 10.23). One unit was defined as one measure for spirits, one glass for red wine/white wine/champagne, or one pint of beer/cider. The raw AC units were transformed by log_2_ (raw AC units + 1) to avoid having a heavily skewed distribution. Smoking intensity was measured in cigarettes per day (CPD) in all current smokers (data-field ID: 3456; *n =* 32,801). Physical activity traits in the UKB were collected from both self-reported questionnaires and devices (wrist-worn accelerometers). METT is a total score of the Metabolic Equivalent Task (MET) minutes per week for walking, moderate activity, and vigorous activity (data-field IDs: 864, 874, 884, 894, 904, and 914; *n =* 41, 938). IPAQ is a derived categorical trait that utilizes the information from the METT and its three subsets mentioned above (see transformation criteria via the link in the **URLs**). The three IPAQ categories are denoted as low, moderate, and high (*n =* 100,611, 190,056, and 127,271). OAA (overall acceleration average) is an objective assessment of physical activity using a wrist-worn accelerometer (data-field ID: 90012; *n* = 97,006). The participants were voluntary, and the measurements were collected for seven days (see Doherty et al.^52^ for more details).

We extracted the phenotypic data of common diseases from the UKB following the disease definitions used in Zhu et al.^43^. There were 22 common diseases in total, and we filtered out 4 diseases with a prevalence < 3% in the UKB. The mean disease count was 1.45 (*s*.*d*. = 1.56) in the whole sample and 1.41 (*s*.*d*. = 1.53) in current drinkers. Body mass index (BMI) was obtained from the physical measurements (data-field: 21001). Educational attainment was indexed by years of school derived from qualification data (data-field ID: 6138). For quantitative traits, extreme phenotypic values outside the mean ± 7 *s*.*d*. range in each sex group were excluded (see below for a more stringent QC step to remove phenotypic outliers after standardisation).

### Correcting for misreports and longitudinal changes

Our MLC corrections consist of two steps. The first step is a phenotypic quality control (QC) procedure used as an attempt to minimize the effects of misreports. We removed the individuals who self-reported as 1) never drinkers (*n =* 14,488), 2) current drinkers with a reported weekly consumption of zero (*n =* 9,064), and 3) current drinkers who provided no response to AC (*n =* 66,058), and retained a total of 349,385 individuals. The second step is to account for self-reported longitudinal changes compared to 10 years ago (data-field ID: 1628). We partitioned the individuals who passed the QC above into three groups based on the longitudinal change (i.e., LESS, SAME or MORE) and conducted GWAS within each group. In the LESS group, we further removed individuals who reduced their AC because of being ill or doctor’s advice, i.e., longitudinal change due to disease ascertainment (data-field ID: 2664; *n =* 11,886). We then performed an inverse-variance weighted meta-analysis of the GWAS results from the three groups (*n =* 336,469). This partitioning strategy efficiently removed any difference in mean or variance between the three groups. More details of the MLC corrections are shown in **Supplementary Figures 1** and **2**.

### Genome-wide association analysis

The UKB genotype data were cleaned and imputed into the Haplotype Reference Consortium (HRC)^53^ panel by the UKB team^23^. We selected a subset of the sample of European ancestry (*n =* 456,426) from the whole UKB cohort by projecting the individuals onto the PCs from the 1000 Genome Project (1KGP). Genotype posterior probabilities were converted to hard-call genotypes using PLINK2 (--hard-call-thresh 0.1)^54^. We removed SNPs with a minor allele count < 5, Hardy-Weinberg equilibrium test *P*-value < 1×10^−6^, missing genotype rate > 5%, or imputation info score < 0.3. For binary traits, we performed BOLT-LMM analysis^55^ with sex, age and the first 10 PCs fitted as covariates and then transformed the estimates of SNP effects on the observed 0-1 scale to odds ratios (OR) by LMOR^56^. For quantitative traits, we adjusted the phenotypic values for sex and age, standardised the adjusted phenotypes to z-scores, excluded individuals with |z| 5, and conducted the BOLT-LMM analysis^55^ with the first 10 PCs as fitted as covariates in the model.

Considering a loss of power due to decreased sample size by MLC corrections, we randomly down-sampled the GWAS data by 21,940 individuals and repeated this process 30 times. We used a z-statistic to test if the number of loci that became non-significant (or changed from non-significant to significant) after the MLC corrections is significantly different from that expected by random down-sampling. The average number of loci that became non-significant due to down-sampling was 10.03 (standard error of the mean (*s. e. m*.) *=* 0.85), significantly (*P =* 2.08 × 10^−12^) smaller than the decrease in the number of genome-wide significant loci due to the MLC corrections (i.e., 16). For the GWAS signals that lost due to down-sampling, the average proportion of significant associations with the metabolic/cardiovascular traits in PheWAS was 31.2% (*s. e. m. =* 3.8), which was significantly lower than the observed 44.9% (*P =* 3.61 × 10^−4^), supporting the enrichment of the 16 loci in metabolic/cardiovascular traits (**Supplementary Table 3**). We also identified 10 loci that became genome-wide significant only after the MLC corrections (**Supplementary Table 2**). The down-sampling analysis showed that only 3.27 loci (*s. e. m. =* 0.52) would be expected by chance (**Supplementary Table 3**), indicating that most of the 10 loci were likely to be masked by MLC in the uncorrected GWAS.

### Estimating heritability and genetic correlation

We used the LD score regression^57^ (LDSC) to estimate SNP-based heritability for a trait and the bivariate-LDSC^35^ to estimate genetic correlation between traits using ~1.2 million SNPs in common with those in HapMap 3 (Ref^58^). For the 234 traits for which we obtained GWAS summary data from LD Hub (**URLs**), the LDSC analyses was performed online in LD Hub^36^. Note that due to the restricted access to the full summary statistics of the 23andMe data sets, we did not perform the genetic correlation analysis for AC using the full GSCAN data^5^.

### Mendelian randomization analysis

Mendelian randomization (MR) is a method to estimate causative effect of an exposure on an outcome using instrumental variables (IVs) associated with the exposure^39,40^. MR assumes that the IVs are independent of possible confounders that may associate with both the exposure and outcome. Also, the IVs are assumed not to be associated with the outcome other than mediated through the exposure. However, in real data, these assumptions can be violated, leading to a biased estimate of the causal effect^59^. We performed MR analyses to test the causal effect of AC on BMI using IVW, MR-Egger and weighted median implemented in the R package ‘MendelianRandomization’ (**URLs**), MR-PRESSO in R and GSMR implemented in GCTA (v1.91.8beta) (**URLs**). The IVs were selected from a clumping analysis of the GWAS summary statistics in GCTA-GSMR (clumping criteria: window size = 1 Mb, *P =* 5 × 10^−8^ and LD *r*^2^ *=* 0.01).

### Simulating data with disease ascertainment

We carried out simulations to mimic the bias due to disease ascertainment in GWAS and its follow-up analyses. If individuals who are affected by a disease tend to change a behaviour, such a change of behaviour would lead to a spurious correlation between the disease and behaviour. We considered four scenarios in the simulation: I) the disease liability (D) is independent of the behavioural trait (Y), and 100 SNPs are associated with Y only; II) Y had a causal effect on D, and 100 SNPs are associated with Y (and D mediated through Y); III) Y and D are independent, and 100 SNPs are associated with D only; IV) Y had a causal effect on D, 100 SNPs affected Y (and D mediated through Y), and another set of 100 SNPs affected D directly. In each scenario, to mimic the disease ascertainment, we reduced the values of Y for the individuals who had high values of D. More specifically, if the D value of an individual passed a threshold (e.g., top 10%), the corresponding Y value would be subtracted by a constant. We set the disease ascertainment threshold to be 10%, 20%, 30%, or 40% and considered a subtraction from Y value by 1, 2, 3, 4 or 5 standard deviations. We then conducted GWAS and estimated SNP effect correlation between Y and D using the *r*_b_ approach developed in a previous study^60^ as well as the causal effect of Y on D using GSMR. To demonstrate the effectiveness of our MLC corrections, we performed an adjustment for longitudinal change in our simulation setting under scenario IV. We divided the individuals into two groups (LESS and SAME) and then conducted the GWAS separately, followed by a meta-analysis. Details of the simulation process and parameter specifications can be found in the **Supplementary Note 2**.

## Data Availability

The individual-level genotype and phenotype data are available through formal application to the UK Biobank (http://www.ukbiobank.ac.uk).

## URLs

PheWAS: http://atlas.ctglab.nl/PheWAS

LDSC: https://github.com/bulik/ldsc

LD Hub: http://ldsc.broadinstitute.org/ldhub/

GSMR: http://cnsgenomics.com/software/gsmr/

MR-PRESSO: https://github.com/rondolab/MR-PRESSO

Other MR methods: https://CRAN.R-project.org/package=MendelianRandomization

IPAQ data processing: http://biobank.ndph.ox.ac.uk/showcase/showcase/docs/ipaq_analysis.pdf

## Acknowledgements

We thank Kyoko Watanabe for the assistance in the PheWAS analysis. This research was supported by the Australian Research Council (FT180100186 and FL180100072), and the Australian National Health and Medical Research Council (1107258, 1173790, and 1113400). This study makes use of data from the UK Biobank (project ID: 12505). A full list of acknowledgements can be found in the **Supplementary Note 5**.

## Author contributions

JY and AX conceived the study. JY, AX and JZ designed the experiment. AX performed all the analyses and simulations. LJ contributed to the analysis of the physical activity data. ZZ contributed to the GSMR analysis. PMV and NRW provided critical advice in data analysis and interpretation of the results. AX, JY and JZ wrote the manuscript with the participation of all authors. All the authors approved the final version of the manuscript.

## Competing interests

The authors declare no competing financial interests.

## Notes

### Competing Interest Statement

The authors have declared no competing interest.

### Author Declarations

This study is approved by the University of Queensland Human Research Ethics Committee (approval number: 2011001173).

